# Development and Evaluation of a Digital Scribe: Conversation Summarization Pipeline for Emergency Department Counseling Sessions towards Reducing Documentation Burden

**DOI:** 10.1101/2023.12.06.23299573

**Authors:** Emre Sezgin, Joseph Sirrianni, Kelly Kranz

**Affiliations:** Nationwide Children’s Hospital, Columbus OH; Ohio State University College of Medicine, Columbus OH

**Keywords:** Text Summarization, Emergency department, clinical conversation, pre-trained language model, Natural Language Processing, documentation burden

## Abstract

**Objective:** We present a proof-of-concept digital scribe system as an ED clinical conversation summarization pipeline and report its performance.

**Materials and Methods:** We use four pre-trained large language models to establish the digital scribe system: T5-small, T5-base, PEGASUS-PubMed, and BART-Large-CNN via zero-shot and fine-tuning approaches. Our dataset includes 100 referral conversations among ED clinicians and medical records. We report the ROUGE-1, ROUGE-2, and ROUGE-L to compare model performance. In addition, we annotated transcriptions to assess the quality of generated summaries.

**Results:** The fine-tuned BART-Large-CNN model demonstrates greater performance in summarization tasks with the highest ROUGE scores (F1_ROUGE-1_=0.49, F1_ROUGE-2_=0.23, F1_ROUGE-L_=0.35) scores. In contrast, PEGASUS-PubMed lags notably (F1_ROUGE-1_=0.28, F1_ROUGE-2_=0.11, F1_ROUGE-L_=0.22). BART-Large-CNN’s performance decreases by more than 50% with the zero-shot approach. Annotations show that BART-Large-CNN performs 71.4% recall in identifying key information and a 67.7% accuracy rate.

**Discussion:** The BART-Large-CNN model demonstrates a high level of understanding of clinical dialogue structure, indicated by its performance with and without fine-tuning. Despite some instances of high recall, there is variability in the model’s performance, particularly in achieving consistent correctness, suggesting room for refinement. The model’s recall ability varies across different information categories.

**Conclusion:** The study provides evidence towards the potential of AI-assisted tools in reducing clinical documentation burden. Future work is suggested on expanding the research scope with larger language models, and comparative analysis to measure documentation efforts and time.

## Introduction

Healthcare professionals (HCPs), including clinicians, nurses, therapists, and other practitioners, dedicate a considerable amount of their working hours to charting and maintaining clinical documentation.[1–3] This labor-intensive process has been linked to burnout among these providers, manifesting as emotional exhaustion, decreased focus, and heightened cognitive burden.[1,2] This issue is particularly prevalent within emergency departments (ED),[4,5] where ED crowding impacts the process due to the high volume of patients waiting to be seen, and low throughput due to limited space, resources, staff and inefficient flow further contributing to delays in treating patients.[6,7] In addition, the use of EMRs has significantly impacted clinical documentation workflow and communication within routine healthcare, influenced by Meaningful Use (MU) requirements, the Affordable Care Act (ACA) reimbursement models, and a heavily regulated environment.[8,9] Literature reported that clinicians spend more time on electronic documentation and administrative tasks than providing direct patient care.[3,10] Clinicians may allocate over half of their working hours to clinical documentation and charting, which has led to decreased direct patient interaction.[11,12] In some cases, insufficient time for documentation leads to burnout.[12] Overall, unaddressed needs and burden may influence unintended choices, as some clinicians express a willingness to remain non-compliant to reduce the burden associated with documentation.[13]

In 2022, the Surgeon General issued an advisory on addressing burnout, which includes several recommendations to address the burden on HCPs in the United States.[14] Some of the recommendations emphasize “designing technology to serve the needs of health workers, care teams, and patients across the continuum of care” and “improving our understanding of how to develop and apply health information technology that more effectively supports health workers in the delivery of care.”[14] In line with that, AMIA 25x5 Task Force issued a call for action to implement personalized clinical decision support (CDS) to improve user-specific workflows and support care recommendations[15] as well as emphasized artificial intelligence as part of current and emerging applications to reduce documentation burden in the long term.[16]

Clinical documentation could be an AI-assisted process, interactively assisting HCPs and easing the burden.[17–19] A *digital scribe* is an “automated clinical documentation system” to capture the HCP conversations with patients and/or other providers and create clinical documentation similar to a human medical scribe [1]. There are several emerging natural language processing (NLP) and deep learning models being used as automated text summarization (ATS) and conversation summarization in the literature [20]. Yet, the implementation of digital scribing in medical informatics and health services has been limited due to technical and algorithmic challenges and limited dataset availability. [1] In this study, we address this gap and present and evaluate a proof-of-concept digital scribe system (as an automated text summarization pipeline) for clinical conversations. We report its performance, with a specific focus on ED consultation sessions.

## Background

ATS is the foundation of the digital scribe, and it aims to automatically generate a concise and clear summary of a text, highlighting the key information for the intended audience.[21–23] ATS can be broadly categorized into two approaches: extractive summarization and abstractive summarization. Extractive summarization selects and combines important sentences and fragments from the original text to form a summary.[24,25] Abstractive summarization (ABS) generates new summaries that incorporate the essential elements of the original text, potentially including key phrases.[24,26] ABS requires both identifying the important aspects of the original text and producing relevant and new natural language summaries.[27] In this study, we used the ABS approach.

Deep learning has been the predominant method for state-of-the-art ABS.[21,28] With the recent development of Transformer network models and the larger generalized language models,[29,30] fine-tuning and/or modifying pre-trained transformer-based models have become the leading techniques for ABS on public datasets.[21] Specialized transformer models have been developed for ABS, such as PEGASUS family of pre-trained models,[21] BART, [31] and its modifications,[32] and T5 family.[33,34] ABS in the biomedical field has mostly focused on online biomedical texts over clinical applications. Overall, ATS has been understudied with medical records as only 11 of the 58 reviewed studies (19%) used Electronic Medical Record (EMR) information as input.[35] However, a recent survey on dialog summarization found that pre-trained language model-based method achieved the highest scores in summarization of public datasets on meeting conversations and chatlogs.[36]

## Methods

### Study setting and data collection

In the scope of our study, we use a dataset (phone conversations) available at Nationwide Children’s Hospital (NCH) Physician Consult and Transfer Center (PCTC).[37] PCTC is a call service that receives calls from healthcare providers across the U.S. to consult, admit, transfer, or refer patients. A nurse team responds to the calls from physicians, registers their calls, connects them to physicians at NCH, and takes a summary note of the conversation into the corresponding patient records (Epic EMR system).[38] Emergency department (ED) patient transfer calls constitute a large amount of the daily PCTC calls. Our proposed digital scribe system uses the conversational data (audio files) stored at NCH servers. Study is approved by NCH ethical board (#00002897)

In this study, 100 phone call recordings from 100 unique callers (physicians) for ED referrals at NCH are used (∼412 total minutes). The calls are randomly selected from the local server (between November-December 2022). Each call consists of a multi-turn conversation (ranging from 1 to 9 minutes conversation each) among PCTC nurses, an ED clinician or staff, and an external clinician or nurse. **Figure 1** outlines the clinical flow and study design.

**Figure 1.**
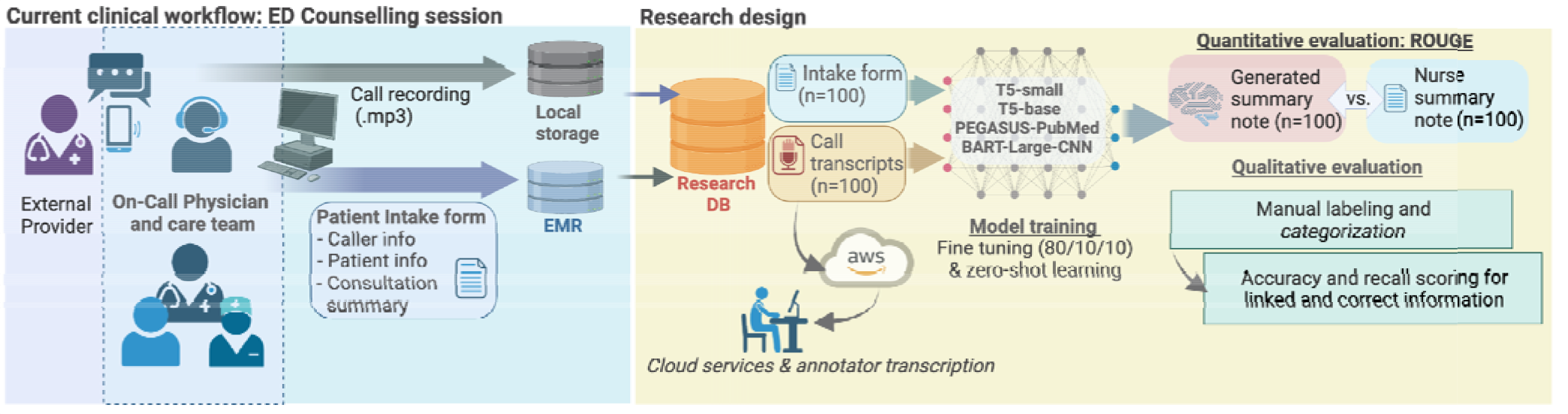
Study design

### Audio transcription

To convert the audio recordings into text, we follow a two-step approach. First, we use speech-to-text services via Amazon Web Services (AWS Transcribe),[39] and then an annotator reviews the original recordings and corrects any errors in the transcript to generate clean transcripts. Dialog between speakers is differentiated with a speaker label (e.g. “Speaker 1: Hello.”). The models have a maximum input token size of 1024 tokens. Of the 100 transcripts, 82 of the transcripts have fewer than 1024 tokens, and the maximum length of the transcript is 1987 tokens (**Figure 2**). Longer transcripts were truncated to include only the first 1024 tokens.

**Figure 2.**
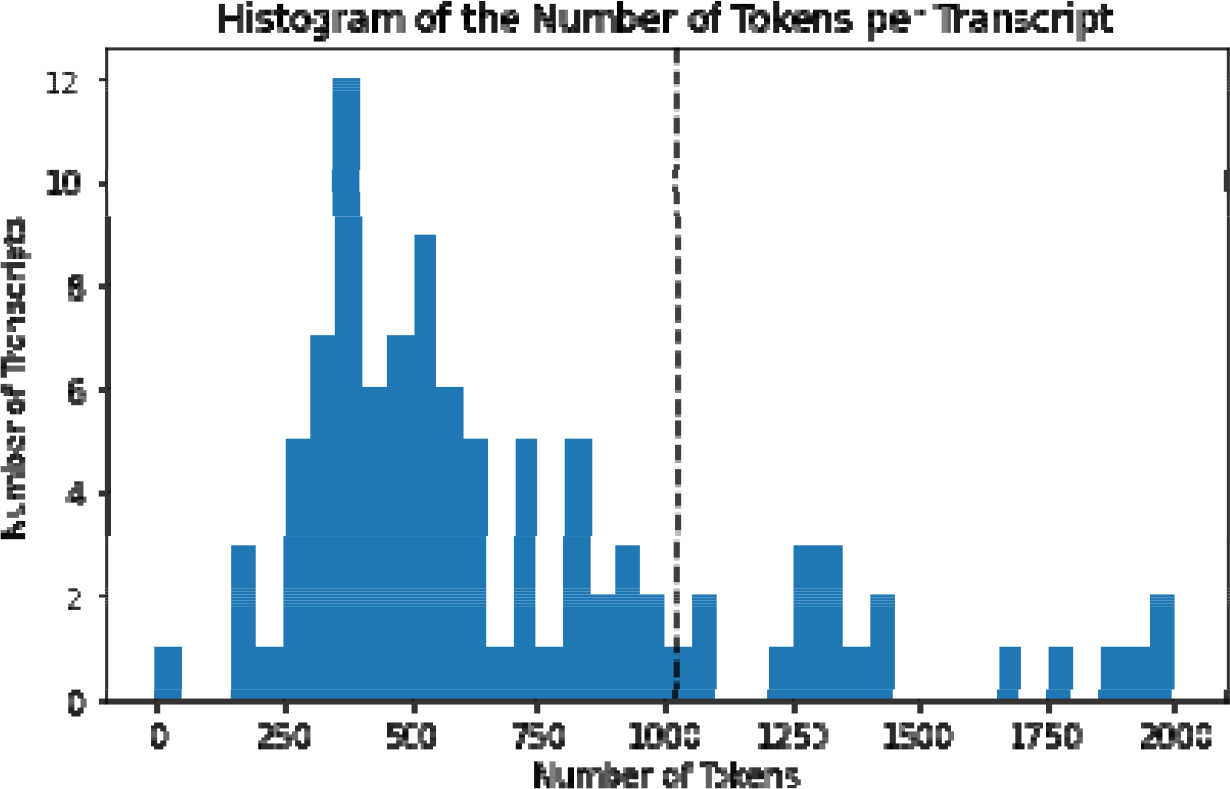
A histogram of the number of tokens per transcript. The tokens were generated for this graph using the BART tokenizer.[40] The vertical line represents the maximum input length of the models, 1024 tokens, and 82% of transcripts clusters to the left of this line.

### Model selection

We employ four pre-trained large language models (T5-small[33], T5-base[33], PEGASUS-PubMed[41], and BART-Large-CNN[40]) for the task of summarizing clinical conversation transcriptions based on their unique strengths and adaptability to the healthcare domain. Our two T5 models use the original T5 seq2seq architecture,[33] trained for a small model (60 million parameters) and a base model (220 million parameters). The T5 models were trained on a large corpus of English text and performed well in tasks like summarization, question answering, and translation. PEGASUS-PubMed (568 million parameters) comes from the class of PEGASUS models[41] developed for abstractive summarization. The inclusion of PEGASUS-PubMed in our selection is driven by its specialization in the biomedical field (Pre-trained in biomedical literature via PubMed repository).[41] BART-Large-CNN (406 million parameters) is a BART model that is fine-tuned on the CNN Daily Mail dataset for summarization. BART-Large-CNN is chosen for its demonstrated effectiveness in producing coherent and contextually accurate summaries.[40]

Our choice of these models is influenced by their combined efficiency, domain-specific accuracy, and ability to produce coherent, reliable summaries, which are critical in the fast-paced and precision-oriented context of healthcare. In addition, these models offer a practical solution, enabling us to process conversation transcriptions quickly without overextending our hardware capabilities (All models were run on a single A100 NVIDIA GPU with 40GB of VRAM), which may represent common computational resources in healthcare.[42,43] Furthermore, our decision is influenced by security, privacy, and compliance. Larger and more resource-intensive LLMs require API access via cloud services. At the time this study was conducted, our team did not have compliant service access to use such models (e.g. GPT, LLaMA) with our dataset which includes Protected Health Information (PHI) and patient data.

### Model training

We use zero-shot (no fine-tuning) and fine-tuning approaches. For fine-tuning, each model is fine-tuned using 10-fold cross-validation (90 training samples, 10 hold-out testing samples for each fold). The final evaluation is run over the concatenated hold-out testing samples from the 10 trials (representing all the data). Each sequence is trained for 30 epochs, with an early stopping patience of 3 epochs, using the AdamW optimizer.[44] Multiple initial learning rates are undertaken (5x10^-10^, 1x10^-6^, 1x10^-5^,1x10^-4^,1x10^-3^, 1x10^-2^) and the best result is reported. For zero-shot, each model is run without any fine-tuning. For training and prediction, each model is configured to use a maximum of 1024 tokens inputs and output up to 200 summary tokens.

The input data (100 transcribed conversations) is summarized and compared with the PCTC nurse notes on each patient’s medical records (structured as details of the complaint, background Information, and consultation recommendations).

### Evaluation

We follow 2-stage evaluation: 1) quantitative evaluation and 2) qualitative evaluation. We report the ROUGE-1, ROUGE-2, and ROUGE-L to compare model performance[45]. ROUGE scores are a standard set of metrics for quantitatively evaluating the similarity of two texts based on the number of common words or word sequences. We compare the summaries generated by each model against the nurse summary notes (ground truth). For this task, we pulled nurse notes from the patient EMR intake form corresponding to each ED referral conversation. We report ROUGE-1 (overlap scores for each word), ROUGE-2 (overlap scores for each bigram), and ROUGE-L (longest common subsequence score).

In addition, we qualitatively evaluate and compare generated summaries against nurse notes to assess the information included in the generated summary. We only evaluate the generated summaries from the best-performing model based on the ROUGE scores. For this qualitative assessment, we compare the amount and type of important information in the nurse notes that is also included in the generated summary. We manually label the nurse notes and generated summaries with the following eight tags: (1)Condition– Symptoms, Diagnosis, Medications related to the patient, (2)Behaviors– The patient’s actions, (3)Measurements– Any numerical value measured, (4)Supplies– List of supplies that the patient has/needs, (5)Date/Time– Any mentioned relevant date or time, (6)Test– Any tests given or not to the patient, (7)Location– Any locations mentioned including where the patient should be brought, (8)Transportation– Method of transportation for the patient.

We incorporate a two-tier annotation system to evaluate the quality of the generated summaries. Firstly, we use *Entity Linking (LINK) annotations* to identify and connect specific pieces of clinical information found in the generated summaries with their corresponding references in the nurse notes. These LINK annotations serve to establish a direct correspondence between the generated text and the ground truth provided by the nurse notes. Secondly, we assess the *Information Accuracy (CORRECT)* of these entity links (LINK).

Information Accuracy is measured by evaluating whether the linked information in the generated summary retains the same meaning as it does in the nurse notes. For instance, if both the nurse and generated summaries report a positive COVID test result for a patient, the LINK is labeled as CORRECT. Conversely, if the generated summary erroneously reports a negative result, the LINK is marked as INCORRECT. This dual-annotation approach allows us to measure not only the presence of key information in the generated summaries but also the accuracy with which it reflects the original nurse notes. The entire process of annotation is facilitated by the use of the MedTator text annotation tool.[37]

## Results

### Quantitative results

Across ROUGE-1 scores, the BART-Large-CNN model displays the highest precision (0.42, CI [0.34, 0.49]), recall (0.53, CI [0.44, 0.62]), and F1-score (0.49, CI [0.38, 0.51]), indicating a strong ability to capture unigrams from the source text (**Table 1**). The T5-base model follows closely, with a ROUGE-1 precision of 0.41 (CI [0.30, 0.51]) and recall of 0.41 (CI [0.32, 0.50]), but a slightly lower F1-score of 0.37 (CI [0.30, 0.45]), suggesting comparable performance in identifying key unigrams. The T5-small and PEGASUS-PubMed models show lower performance on these metrics, with the PEGASUS-PubMed model exhibiting the lowest F1-score of 0.28 (CI [0.22, 0.36]). Similar to ROUGE-1 scores, BART-Large-CNN has the highest Recall (ROUGE-2=0.28, ROUGE-L=0.43) and F1-scores (ROUGE-2=0.23, ROUGE-L=0.35), while T5-base has the highest Precisions scores (ROUGE-2=0.22, ROUGE-L=0.34).

**Table 1.** ROUGE-1, -2, and -L average precision, recall, and F1 scores for the fine-tuned models on clean transcripts (CI= 95% Confidence interval)

| Model | ROUGE-1 Scores |  |  |
| --- | --- | --- | --- |
|  | Precision (CI) | Recall (CI) | F1-score (CI) |
| T5-small | 0.34 (0.26, 0.43) | 0.40 (0.31, 0.50) | 0.35 (0.28, 0.42) |
| T5-base | 0.41 (0.30, 0.51) | 0.41 (0.32, 0.50) | 0.37 (0.30, 0.45) |
| PEGASUS-PubMed | 0.29 (0.21, 0.38) | 0.35 (0.26, 0.44) | 0.28 (0.22, 0.36) |
| BART-Large-CNN | 0.42 (0.34, 0.49) | 0.53 (0.44, 0.62) | 0.49 (0.38, 0.51) |
| Model | ROUGE-2 Scores |  |  |
|  | Precision (CI) | Recall (CI) | F1-score (CI) |
| T5-small | 0.17 (0.13, 0.32) | 0.21 (0.15, 0.29) | 0.18 (0.13, 0.23) |
| T5-base | 0.22 (0.15, 0.30) | 0.22 (0.15, 0.30) | 0.20 (0.15, 0.26) |
| PEGASUS-PubMed | 0.11 (0.07, 0.16) | 0.14 (0.09, 0.20) | 0.11 (0.07, 0.16) |
| BART-Large-CNN | 0.21 (0.16, 0.27) | 0.28 (0.21, 0.36) | 0.23 (0.18, 0.29) |
| Model | ROUGE-L Scores |  |  |
|  | Precision (CI) | Recall (CI) | F1-score (CI) |
| T5-small | 0.28 (0.22, 0.35) | 0.34 (0.25, 0.43) | 0.29 (0.23, 0.35) |
| T5-base | 0.34 (0.25, 0.44) | 0.34 (0.27, 0.44) | 0.32 (0.25, 0.39) |
| PEGASUS-PubMed | 0.22 (0.16, 0.30) | 0.27 (0.20, 0.30) | 0.22 (0.16, 0.29) |
| BART-Large-CNN | 0.33 (0.27, 0.41) | 0.43 (0.34, 0.52) | 0.35 (0.29, 0.42) |

Table 2 reports the performance of the zero-shot models. For ROUGE-1 scores, BART-Large-CNN exhibits the highest precision (0.26, CI [0.19, 0.34]) and recall (0.23, CI [0.17, 0.30]), with a corresponding F1-score of 0.23 (CI [0.17, 0.29]), suggesting a modest capability to identify key unigrams without fine-tuning. The T5-base model also shows relatively better performance compared to T5-small, with precision, recall, and F1-score of 0.30 (CI [0.22, 0.38]), 0.17 (CI [0.15, 0.23]), and 0.20 (CI [0.15, 0.26]), respectively. T5-small has lower scores, and PEGASUS-PubMed’s performance is notably minimal, with an F1-score of 0.07 (CI [0.05, 0.10]). When examining ROUGE-2 scores, which evaluate bigram overlap, the models perform generally poorly, with BART-Large-CNN leading at a lower precision of 0.08 (CI [0.04, 0.12]) and a corresponding F1-score of 0.07 (CI [0.04, 0.10]). The T5 models report low scores, with T5-base obtaining an F1-score of 0.06 (CI [0.03, 0.09]), marginally outperforming T5-small, which has an F1 of 0.05 (CI [0.02, 0.09]). PEGASUS-PubMed has no bigram overlap in this scenario, reflecting significant limitations in its zero-shot performance. Regarding the ROUGE-L scores, BART-Large-CNN achieves the highest F1-score of 0.16 (CI [0.12, 0.21]), albeit modest, indicating its relative advantage in capturing the longest common subsequences in the zero-shot learning context. T5-base and T5-small achieve F1-scores of 0.15 (CI [0.11, 0.21]) and 0.13 (CI [0.08, 0.17]), respectively, followed by PEGASUS-PubMed with an F1-score of 0.06 (CI [0.04, 0.07]).

### Qualitative Results

We label each of the 100 ground truth summaries and the summaries generated by the Bart-Large-CNN model (fine-tuned on 90 not held-out data for that cross-validation fold) using eight tag categories: Conditions, Behaviors, Measurements, Supplies, Date/Time, Tests, Locations, and Transportation.

**Table 2.** ROUGE-1, -2, and -L average precision, recall, and F1 scores for the zero-shot models on clean transcripts (CI= 95% Confidence interval)

| Model | ROUGE-1 Scores |  |  |
| --- | --- | --- | --- |
|  | Precision (CI) | Recall (CI) | F1-score (CI) |
| T5-small | 0.24 (0.17, 0.32) | 0.15 (0.11, 0.22) | 0.17 (0.11, 0.24) |
| T5-base | 0.30 (0.22, 0.38) | 0.17 (0.15, 0.23) | 0.20 (0.15, 0.26) |
| PEGASUS-PubMed | 0.06 (0.04, 0.09) | 0.12 (0.05, 0.16) | 0.07 (0.05, 0.10) |
| BART-Large-CNN | 0.26 (0.19, 0.34) | 0.23 (0.17, 0.30) | 0.23 (0.17, 0.29) |
| Model | ROUGE-2 Scores |  |  |
|  | Precision (CI) | Recall (CI) | F1-score (CI) |
| T5-small | 0.06 (0.02, 0.11) | 0.04 (0.01, 0.08) | 0.05 (0.02, 0.09) |
| T5-base | 0.08 (0.04, 0.12) | 0.05 (0.02, 0.08) | 0.06 (0.03, 0.09) |
| PEGASUS-PubMed | 0.00 (0.00, 0.00) | 0.00 (0.00, 0.00) | 0.00 (0.00, 0.01) |
| BART-Large-CNN | 0.08 (0.04, 0.12) | 0.07 (0.03, 0.11) | 0.07 (0.04, 0.10) |
| Model | ROUGE-L Scores |  |  |
|  | Precision (CI) | Recall (CI) | F1-score (CI) |
| T5-small | 0.18 (0.12, 0.23) | 0.11 (0.07, 0.16) | 0.13 (0.08, 0.17) |
| T5-base | 0.21 (0.16, 0.26) | 0.12 (0.08, 0.21) | 0.15 (0.11, 0.21) |
| PEGASUS-PubMed | 0.05 (0.03, 0.06) | 0.09 (0.06, 0.12) | 0.06 (0.04, 0.07) |
| BART-Large-CNN | 0.18 (0.13, 0.24) | 0.16 (0.11, 0.22) | 0.16 (0.12, 0.21) |

\* CI= 95% Confidence interval

Table 3 presents the average recall for manually annotated information tags in summaries of the fine-tuned BART-Large-CNN. All summaries contain at least one of the specified tags, with an average of 8.67 tags per summary. When examining the average LINK recall, the model performs consistently, with a mean recall of 0.71 (SD=0.23), indicating that over 70% of the information present in the ground truth summaries is also found in the generated summaries. The average CORRECT recall is marginally lower at 0.67 (SD=0.23), suggesting that while the model is proficient at identifying relevant information, there is a slight decrease in accuracy when considering the correctness of the information. **Figure 3** illustrates the recall characteristics of the fine-tuned BART-Large-CNN model.

**Figure 3.**
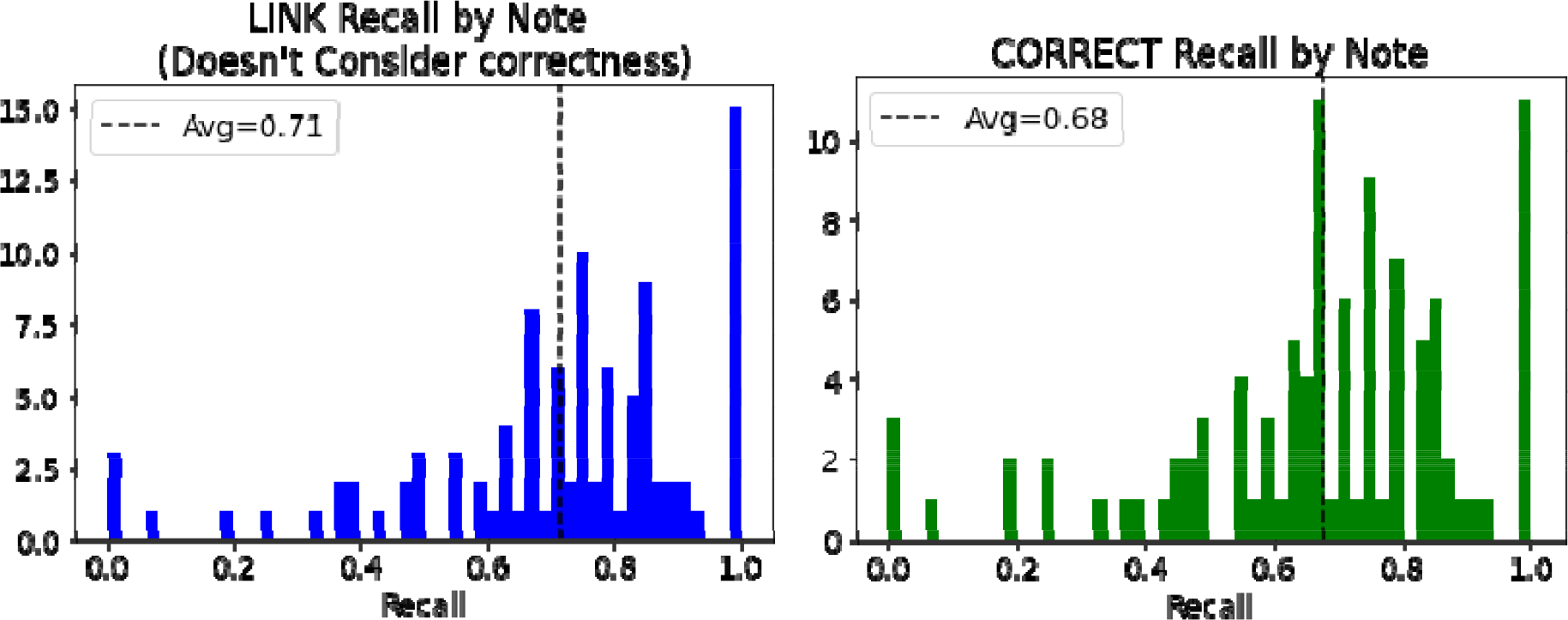
Histograms showing information recalled (without consideration of correctness) [right] and correctly recalled information [left] by a generated summary that appeared in the ground truth summary.

The ‘Condition’ tag appears in 99% (99/100) of the summaries, and it has a high CORRECT recall at 0.73 (SD=0.27), which indicates a high degree of precision in reporting patient conditions, symptoms, and diagnoses. However, tags such as ‘Transportation’ are present in only 41% (41/100) of the summaries, with the lowest average LINK and CORRECT recall scores of 0.44 (SD=0.5). ‘Behaviors’ and ‘Supplies’ tags appear less frequently at 29% (29/100) and 7% (7/100) respectively, yet show relatively high CORRECT recall. **Figure 4** shows an example note sample outlining CORRECT and LINK annotations and tags.

**Table 3.** Average Recall for Tags and Annotations in Generated Summaries by the Fine-tuned BART-Large-CNN Model (SD: Standard Deviation)

| Tags | % Summary with at least 1 Tag | Average Tags per Summary (SD) | Average LINK Recall per Summary (SD) | Average CORRECT Recall per Summary (SD) |
| --- | --- | --- | --- | --- |
| All Tags | 100% (100/100) | 8.670 (4.800) | 0.714 (0.231) | 0.677 (0.228) |
| Condition | 99% (99/100) | 4.848 (2.776) | 0.744 (0.268) | 0.731 (0.274) |
| Behaviors | 29% (29/100) | 1.483 (1.038) | 0.772 (0.380) | 0.772 (0.380) |
| Measurements | 47% (47/100) | 2.298 (1.687) | 0.736 (0.409) | 0.644 (0.425) |
| Supplies | 7% (7/100) | 1.143 (0.350) | 0.571 (0.495) | 0.571 (0.495) |
| DateTime | 46% (46/100) | 1.304 (0.655) | 0.741 (0.409) | 0.730 (0.409) |
| Test | 35% (35/100) | 2.343 (1.453) | 0.673 (0.409) | 0.564 (0.423) |
| Location | 42% (42/100) | 1.071 (0.258) | 0.762 (0.426) | 0.667 (0.471) |
| Transportation | 41% (41/100) | 1.000 (0.000) | 0.439 (0.496) | 0.439 (0.496) |

**Figure 4.**
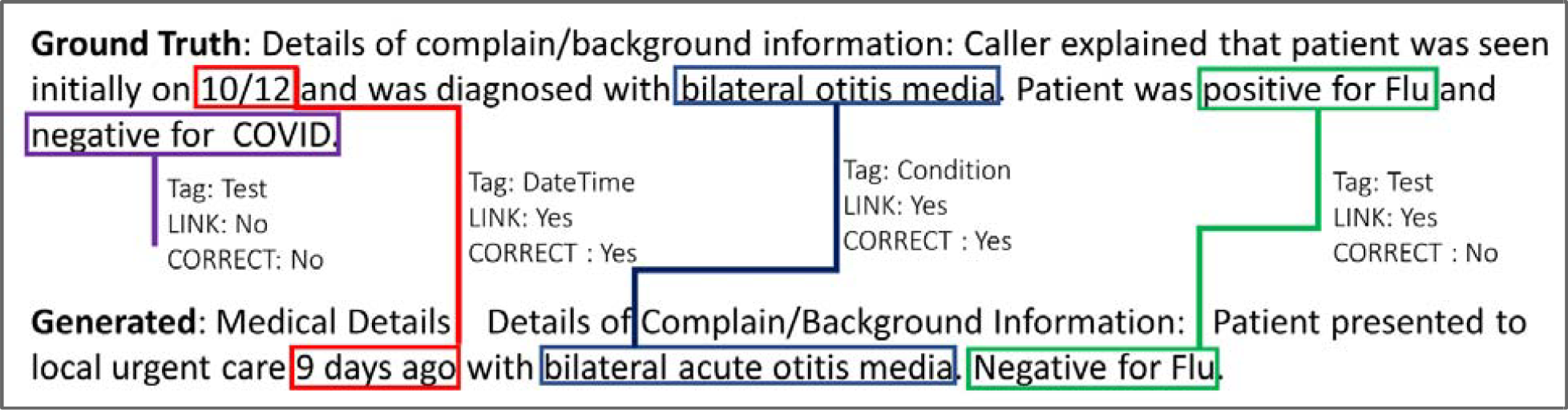
Example generated and nurse note samples with LINK and CORRECT annotations

For all summaries combined, the model demonstrates a LINK recall of 69.7% (604/867) instances where tagged information in the ground truth also appears in the generated summaries (**Table 4**). The CORRECT recall, which indicates the instances where the tagged information from the ground truth summary appears accurately in the generated summary, is slightly lower at 65.7% (570/867). However, of the information that is LINKed correctly, the CORRECT accuracy is high at 94.4% (570/604), indicating that when the model does capture relevant information, it tends to be accurate. ‘Conditions’ shows the highest LINK recall at 72.1% (346/480), and an almost equivalent CORRECT recall at 70.8% (340/480). The CORRECT accuracy for ‘Conditions’ is at 98.3% (340/346), indicating that nearly all the condition-related information captured by the model is accurate. The ‘Behaviors’ and ‘Supplies’ tags have the fewest instances but achieve a CORRECT recall of 74.4% (32/43) and 62.5% (5/8), respectively, with both categories achieving CORRECT accuracy of 100%. Conversely, ‘Test’ and ‘Transportation’ tags display lower performance on LINK and CORRECT recall.

**Table 4.** Information tag appearance and correctness in the summaries generated by the fine-tuned BART-Large-CNN model.

| Tag | Total Tags | LINK recall across all summaries | CORRECT recall across all summaries | CORRECT accuracy across LINKed tags |
| --- | --- | --- | --- | --- |
| All Tags | 867 | 69.7% (604/867) | 65.7% (570/867) | 94.4% (570/604) |
| Condition | 480 | 72.1% (346/480) | 70.8% (340/480) | 98.3% (340/346) |
| Behaviors | 43 | 74.4% (32/43) | 74.4% (32/43) | 100.0% (32/32) |
| Measurements | 108 | 68.5% (74/108) | 57.4% (62/108) | 83.8% (62/74) |
| Supplies | 8 | 62.5% (5/8) | 62.5% (5/8) | 100.0% (5/5) |
| Date/Time | 60 | 75.0% (45/60) | 73.3% (44/60) | 97.8% (44/45) |
| Test | 82 | 62.2% (51/82) | 48.8% (40/82) | 78.4% (40/51) |
| Location | 45 | 73.3% (33/45) | 64.4% (29/45) | 87.9% (29/33) |
| Transportation | 41 | 43.9% (18/41) | 43.9% (18/41) | 100.0% (18/18) |

### Transcription differences

We compare the difference in performance between the AWS transcripts and the clean transcripts. BART-Large-CNN’s ROUGE-1 improves by 0.06 (F1-score) when using the clean transcripts. However, T5-base and PEGASUS-PubMed both have lower F1-scores when using the clean transcripts. This difference is mostly not applicable for ROUGE-2 and ROUGE-L scores with a difference between F1-scores less than 0.02. Please see **Appendix 1** for ROUGE scores of AWS transcripts.

## Discussion

Our fine-tuned text summarization models report promising results compared to similar applications and tasks[20]. The BART-Large-CNN model shows a greater ability to comprehend and replicate the structure and flow of clinical dialogue in medical conversation with a fine-tuned and zero-shot approach. This is similar to the performance of high-performing models on the non-medical CNN/DailyMail dataset.[21] However, the variants of recall show an inconsistency in performance, with a subset of notes being replicated with high accuracy, yet a broader variability indicating room for refinement, especially in achieving consistent correctness. The differential performance across various information categories illuminates the necessity for enhancing model recognition capabilities.[46] As the accuracy rates across most tags are promising, they also highlight the disparity in the model’s ability to uniformly identify and convey the full spectrum of clinically relevant information present in the reference summaries.[47] In a zero-shot context, each model performed relatively worse than their fine-tuned counterparts.

Bart-Large-CNN and T5 have better performance, as the models tend to reproduce some lines of the transcript as the summary. PEGASUS-PubMed, by comparison, outputs similar to the original training data text which is somewhat related to the text in the transcript. These results reinforce the idea that competent zero-shot performance might be achievable at larger model sizes as well as incorporating different architectures and datasets.[48] Furthermore, the variability in model performance in our study, particularly in the context of recall, denotes a significant opportunity for advancing the model’s performance with hybrid models[49] or approaches (e.g., user interface design, human-in-the-loop),[50,51] thereby augmenting its utility in real-world clinical documentation.

### Transcription quality

The transcription quality notably impacts the model performance, as evidenced by the improvement in BART-Large-CNN’s ROUGE-1 scores when utilizing clean transcripts. This improvement underscores the importance of high-quality input data for the efficacy of AI-driven clinical documentation.[52] Interestingly, T5-base and PEGASUS-PubMed models register a lower F1-score with clean transcripts, an anomaly that suggests a complex interaction between model architecture and data quality. This observation requires a closer examination of the preprocessing steps and the models’ resilience to variations in data quality. In the high-stress, fast-paced ED environment, where documentation accuracy is important, these findings highlight the necessity for robust digital scribe systems capable of handling the inherent variability in clinical speech and text data. The minor differences in ROUGE-2 and ROUGE-L scores with different transcript quality suggest that for capturing the broader context and relationships within the text, the models are less sensitive to transcription errors. This resilience is critical for the practical deployment of digital scribes, where they must perform reliably across varying conditions of data quality.[53]

### The nature of conversations

In our observation of audio conversations, we note a common pattern involving additional clinicians or healthcare workers, often leading to multi-participant calls and extended discussions. The conversation starts with caller information and patient information exchange, followed by patient health information shared later in the conversation. Waiting times with hold tones are frequent. A notable discrepancy between audio summaries and intake notes is that, especially when nurses follow up for additional details, these details are not always included in the initial transcription. Another observation is the variation in note style and content, depending on the nurse taking the notes, indicating differences in documentation approaches among nurses. This added an extra layer of complexity to the task of accurate digital scribing.

Additionally, external factors like background noise and coughing during conversations pose potential challenges for automated transcription accuracy.[1] The intake notes sometimes include details from internal consultations not present in the original audio, pointing to a possible mismatch in the documentation. These insights underscore the multifaceted nature of clinical communication and the challenges it presents for effective digital documentation.[54]

### Implications

The implications of our study extend into several key areas of healthcare informatics and policy. Firstly, the use of the BART-Large-CNN model in clinical documentation points towards a potential to reduce the documentation burden on HCPs, aligning with the broader goal of mitigating burnout.[1] The high accuracy in key information categories like ‘Conditions’ indicates that AI-assisted tools can effectively complement HCPs’ condition tasks. However, the successful integration of such AI tools hinges on their design and usability.[55,56] The variability in model performance underscores the need for a user-centered design approach and a systems thinking approach to overcome technical challenges.[57,58] This involves tailoring these tools to fit into clinical workflows, ensuring they are intuitive and capable of handling the dynamic nature of clinical environments.[59]

In line with recommendations by the Surgeon General and the AMIA 25x5 Task Force, the findings inform developing and applying health information technology that supports HCPs, suggesting that policies may encourage the exploration and adoption of AI tools like digital scribes in clinical settings.[15] This could be achieved through incentives for technology adoption, support for implementation research and technical development, and the development of evidence-based guidelines to ensure ethical and secure use of AI in healthcare.[60] However, the collaboration between HCP and AI is key to success in improving the accuracy, consistency, and completeness of medical documentation while minimizing documentation errors.[51,61] It is also important to develop operationalization and implementation plans with accountable, fair, and inclusive AI approaches to ensure the trustworthiness of the digital scribes. [62,63]

### Limitations

The limitations of our study are multifaceted, reflecting both methodological constraints and broader challenges in the field. Firstly, the absence of standardized and validated measures for assessing documentation burden presents a significant challenge.[64] Therefore we depend on our quantitative and qualitative approaches to assess quality, and assuming higher quality of summarization will contribute to reducing documentation burden. Our scoring does not account for differences in notes, note-takers (nurses), and conversations. ROUGE metrics are coherence-insensitive, focusing solely on word overlap without considering the coherence and logical flow of the summaries, which introduces a limitation for quantitative analysis.[65]. Our qualitative evaluation focused on a 2-tier assessment, which might limit the perspectives. The study lacks qualitative feedback from nurses and clinicians to further assess the perceived value and utility of generated summaries. These limitations are compounded by the small dataset size, single annotator bias, lack of real-world testing, and the limited scope of the dataset for ED referrals, all of which contribute to potential constraints on the generalizability and applicability of our findings.

### Future work

In future works, we aim to expand the scope and applicability of our research. A primary focus will be on testing a cloud-based transcription and digital scribe pipeline using advanced language models with larger and diverse datasets. This initiative will be geared towards developing a deployable pipeline, with a specific scenario involving a call service connection and providing immediate feedback through a web application to nurses. Another important area of exploration will be the hybrid models[35] combining statistical, machine learning, and computational linguistics techniques, and experimenting via a comparative study utilizing emerging documentation measures focusing on effort, time, and other relevant units of analysis. [64]

## Conclusions

Our study introduces the development and testing of a digital scribe pipeline, contributing to the field of automated clinical documentation and efficient documentation flow. By utilizing a real-world dataset, our research addresses a critical gap in the literature, particularly in the areas of workflow optimization and clinical and nurse informatics applications [1]. The practical implications of our findings are offering potential time and resource savings for healthcare systems, aiming to reduce the documentation burden among nurses and clinicians, thereby enhancing overall healthcare delivery efficiency and quality.

## Data Availability

All data produced in the present study are available upon ethical board approval and subject to institutional data use agreement.

## Conflict of interest

None declared.

## Funding

The project described was supported by Award Number UM1TR004548 from the National Center for Advancing Translational Sciences. The content is solely the responsibility of the authors and does not necessarily represent the official views of the National Center for Advancing Translational Sciences or the National Institutes of Health.

## Acknowledgment

We acknowledge the contribution of Mark Wang and Daniel Jackson on data cleaning, and Simon Linwood on project ideation. We are thankful to the PCTC team for their feedback. Figure 1 is created using BioRender.com.

## Appendix 1. AWS transcripts ROUGE scores

ROUGE-1 Scores
| Model | Precision (CI) | Recall | F1-score |
| --- | --- | --- | --- |
| T5-small | 0.32 (0.24, 0.40) | 0.40 (0.31, 0.49) | 0.33 (0.26, 0.39) |
| T5-base | 0.42 (0.32, 0.52) | 0.39 (0.30, 0.49) | 0.38 (0.30, 0.45) |
| Pegasus-Pubmed | 0.29 (0.21, 0.38) | 0.36 (0.28, 0.45) | 0.30 (0.23, 0.37) |
| Bart-Large-CNN | 0.40 (0.32, 0.48) | 0.51 (0.43, 0.59) | 0.43 (0.37, 0.48) |

ROUGE-2 Scores
| Model | Precision | Recall | F1-score |
| --- | --- | --- | --- |
| T5-small | 0.16 (0.11, 0.21) | 0.20 (0.14, 0.27) | 0.16 (0.12, 0.21) |
| T5-base | 0.22 (0.16, 0.29) | 0.21 (0.15, 0.29) | 0.20 (0.15, 0.26) |
| Pegasus-Pubmed | 0.11 (0.07, 0.15) | 0.14 (0.09, 0.18) | 0.11 (0.08, 0.15) |
| Bart-Large-CNN | 0.20 (0.15, 0.26) | 0.26 (0.20, 0.34) | 0.22 (0.17, 0.27) |

ROUGE-L Scores
| Model | Precision | Recall | F1-score |
| --- | --- | --- | --- |
| T5-small | 0.26 (0.19, 0.27) | 0.32 (0.24, 0.41) | 0.27 (0.21, 0.32) |
| T5-base | 0.35 (0.26, 0.44) | 0.33 (0.25, 0.42) | 0.31 (0.25, 0.38) |
| Pegasus-Pubmed | 0.22 (0.16, 0.29) | 0.28 (0.21, 0.35) | 0.23 (0.17, 0.29) |
| Bart-Large-CNN | 0.32 (0.25, 0.39) | 0.41 (0.33, 0.50) | 0.34 (0.28, 0.40) |

Zero-shot models - ROUGE scores with AWS transcripts ROUGE-1 Scores
| Model | Precision | Recall | F1-score |
| --- | --- | --- | --- |
| T5-small | 0.18 (0.12, 0.27) | 0.11 (0.06, 0.17) | 0.13 (0.08, 0.19) |
| T5-base | 0.27 (0.20, 0.35) | 0.16 (0.11, 0.22) | 0.19 (0.14, 0.25) |
| Pegasus-Pubmed | 0.07 (0.04, 0.10) | 0.12 (0.09, 0.16) | 0.08 (0.06, 0.11) |
| Bart-Large-CNN | 0.25 (0.18, 0.31) | 0.22 (0.17, 0.28) | 0.22 (0.17, 0.27) |

ROUGE-2 Scores
| Model | Precision | Recall | F1-score |
| --- | --- | --- | --- |
| T5-small | 0.04 (0.01, 0.08) | 0.03 (0.01, 0.05) | 0.03 (0.01, 0.06) |
| T5-base | 0.07 (0.03, 0.11) | 0.04 (0.02, 0.07) | 0.05 (0.02, 0.08) |
| Pegasus-Pubmed | 0.00 (0.00, 0.01) | 0.01 (0.00, 0.01) | 0.00 (0.00, 0.01) |
| Bart-Large-CNN | 0.07 (0.04, 0.10) | 0.06 (0.03, 0.10) | 0.06 (0.03, 0.09) |

ROUGE-L Scores
| Model | Precision | Recall | F1-score |
| --- | --- | --- | --- |
| T5-small | 0.14 (0.09, 0.20) | 0.08 (0.05, 0.12) | 0.10 (0.06, 0.14) |
| T5-base | 0.19 (0.14, 0.25) | 0.12 (0.08, 0.16) | 0.14 (0.10, 0.18) |
| Pegasus-Pubmed | 0.05 (0.03, 0.07) | 0.09 (0.07, 0.13) | 0.06 (0.04, 0.08) |
| Bart-Large-CNN | 0.17 (0.12, 0.21) | 0.16 (0.11, 0.21) | 0.15 (0.11, 0.20) |

## Notes

### Competing Interest Statement

The authors have declared no competing interest.

### Author Declarations

Study is approved by NCH ethical board (#00002897)

